# Feasibility assessment of telehealth-based non-communicable disease screening and monitoring by community health leaders in remote areas: A pilot study in Chiang Mai, Thailand

**DOI:** 10.1101/2025.03.11.25323802

**Authors:** Sujittra Kaewkart, Pitaya Suebtam, Sawittree Thatong, Nisachon Kaewkart, Aphirak Pinasu, Premmarin Inmonthian, Napatsakorn Kohklang, Kittichai Wantanajittikul, Sakorn Pornprasert, Faiz Shah, Tanawan Samleerat Carraway, Woottichai Khamduang

**Affiliations:** Department of Medical Technology, Faculty of Associated Medical Sciences, Chiang Mai University, Chiang Mai, Thailand; LUCENT international collaboration, Faculty of Associated Medical Sciences, Chiang Mai University, Chiang Mai, Thailand; Department of Family Medicine and Primary Care, Lamphun Hospital, Lamphun, Thailand; Surgical Nursing Division, Maharaj Nakorn Chiang Mai Hospital, Faculty of Medicine, Chiang Mai University, Chiang Mai, Thailand; Department of Radiologic Technology, Faculty of Associated Medical Sciences, Chiang Mai University, Chiang Mai, Thailand; Yunus Center AIT, Asian Institute of Technology, Klong Luang, Pathum Thani, Thailand

## Abstract

Limited access to healthcare in remote areas poses a significant challenge for non-communicable disease (NCD) prevention. This study assessed the feasibility and acceptability of *HealthD*, a telehealth platform enabling Community Health Leaders (CHLs) to screen and monitor NCD risks in rural Thailand. From June to November 2023, a prospective study was conducted in Chiang Mai, Thailand, involving 120 adults (aged 30–70 years) with no prior NCD diagnosis. Trained CHLs performed risk assessments for diabetes mellitus (DM), cardiovascular disease (CVD), and chronic obstructive pulmonary disease (COPD) using the *HealthD* system. Participants were categorized into four risk levels and received tailored teleconsultations at two- and four-month follow-ups. Satisfaction surveys were conducted among CHLs and participants. At baseline, 73% of participants were at risk for DM (21% high-risk), 3% for CVD, and 10% for COPD. Over follow-up visits, DM risk fluctuated (63% at two months; 76% at four months), while CVD (4% to 6%) and COPD (6% to 8%) risk levels remained relatively stable. CHLs demonstrated significant competency improvement post-training (mean score increase: 8.4 to 9.3, *p* = 0.010). High satisfaction levels were reported among CHLs (92%) and participants (97%), indicating strong acceptability of the *HealthD* system. The *HealthD* telehealth platform is a feasible and well-accepted solution for community-based NCD screening and monitoring. Its integration into Thailand’s healthcare system could enhance early disease detection and improve health outcomes in remote communities.

## Introduction

Non-communicable diseases (NCDs) pose a major global health challenge, accounting for approximately 74% of global deaths, with cardiovascular diseases (CVDs), diabetes mellitus (DM), chronic obstructive pulmonary disease (COPD), and cancer being the most prevalent conditions [1]. The World Health Organization (WHO) reports that 17 million premature NCD-related deaths occur annually, with a disproportionate burden in low- and middle-income countries (LMICs) [1, 2]. In Thailand, NCDs contribute to 77% of all deaths, with leading causes including cancer (24%), CVDs (23%), diabetes (6%), and chronic respiratory diseases (5%) [2, 3]. In 2022, Chiang Mai province reported 159 NCD-related deaths per 100,000 people, highlighting a growing health crisis [4]. The increasing prevalence of NCDs places significant strain on healthcare systems, leading to hospital overcrowding, longer waiting times, and reduced patient satisfaction [5]. Moreover, chronic conditions result in productivity losses and impose substantial economic burdens, with Thailand’s government allocating approximately 4.1 billion USD annually for NCD management [6]. Despite the establishment of Thailand’s Universal Coverage Scheme (UCS) under the National Health Security Act B.E. 2545 (2002), disparities in healthcare access persist, particularly in rural and remote areas, where geographic and economic barriers limit routine health screenings and timely interventions [7, 8].

To address these challenges, community-based healthcare models and decentralized health systems have been proposed to improve early NCD detection and management [9, 10]. Community Health Leaders (CHLs) play a critical role in bridging the gap between healthcare providers and underserved populations, offering localized health promotion, preventive care, and basic health assessments [11]. The integration of telehealth technology further strengthens this approach by enabling remote health monitoring, real-time consultations, and digital health record-keeping [12]. Telehealth interventions have successfully improved healthcare accessibility in LMICs such as Bangladesh, India, and Indonesia, demonstrating their potential for scalable, cost-effective NCD management [13–18].

In Thailand, telehealth adoption has expanded, particularly during the COVID-19 pandemic, with applications in cancer care, HIV management, telenursing, and telerehabilitation [19–22]. However, the use of telehealth for community-based NCD screening and monitoring remains limited. To address this gap, we developed the *HealthD* system, a telehealth platform designed to empower CHLs in conducting NCD risk assessments and follow-up monitoring in remote Thai communities.

This study evaluates the feasibility and acceptability of the *HealthD* system for NCD screening and monitoring in rural Thailand. Specifically, we assess: 1) the ability of trained CHLs to conduct telehealth-based NCD screenings. 2) the ability of *HealthD*’s risk assessment algorithm in categorizing patients based on NCD risk levels, 3) the acceptability and satisfaction of CHLs and participants with the system. By addressing these objectives, this study goal is to provide evidence for integrating telehealth-based NCD screening into Thailand’s national healthcare programs, potentially improving health outcomes in underserved populations.

## Materials and methods

### Overview of the *HealthD* system

The *HealthD* system was developed to enhance NCD screening and teleconsultation in remote communities by leveraging portable diagnostic tools and digital health technology. At the core of the system is the “*HealthD* backpack”, a fully equipped mobile health kit containing vital sign monitoring devices, physical examination tools, point-of-care testing (POCT) laboratory devices, sanitary and biohazard waste disposal supplies, and a tablet personal computer (tablet PC) for data collection and teleconsultation. The system integrates the T-logic algorithm, a web-based decision-support tool that classifies participants into four NCD risk categories-green (normal), yellow (low risk), orange (moderate risk), and red (high risk)-using international reference standards adapted for the Thai population. Trained Community Health Leaders (CHLs) conducted on-site screenings, collecting participants’ socio-demographic data, vital signs, and NCD risk assessment results (excluding cancer markers). These data were securely uploaded to an online database, where they were analyzed and categorized by the T-logic algorithm. Participants were then provided with risk-specific teleconsultations, with follow-up assessments conducted at two and four months. At the study’s conclusion, data analysis and participant satisfaction surveys were performed to evaluate the system’s effectiveness and acceptability, Fig 1

**Fig 1.**
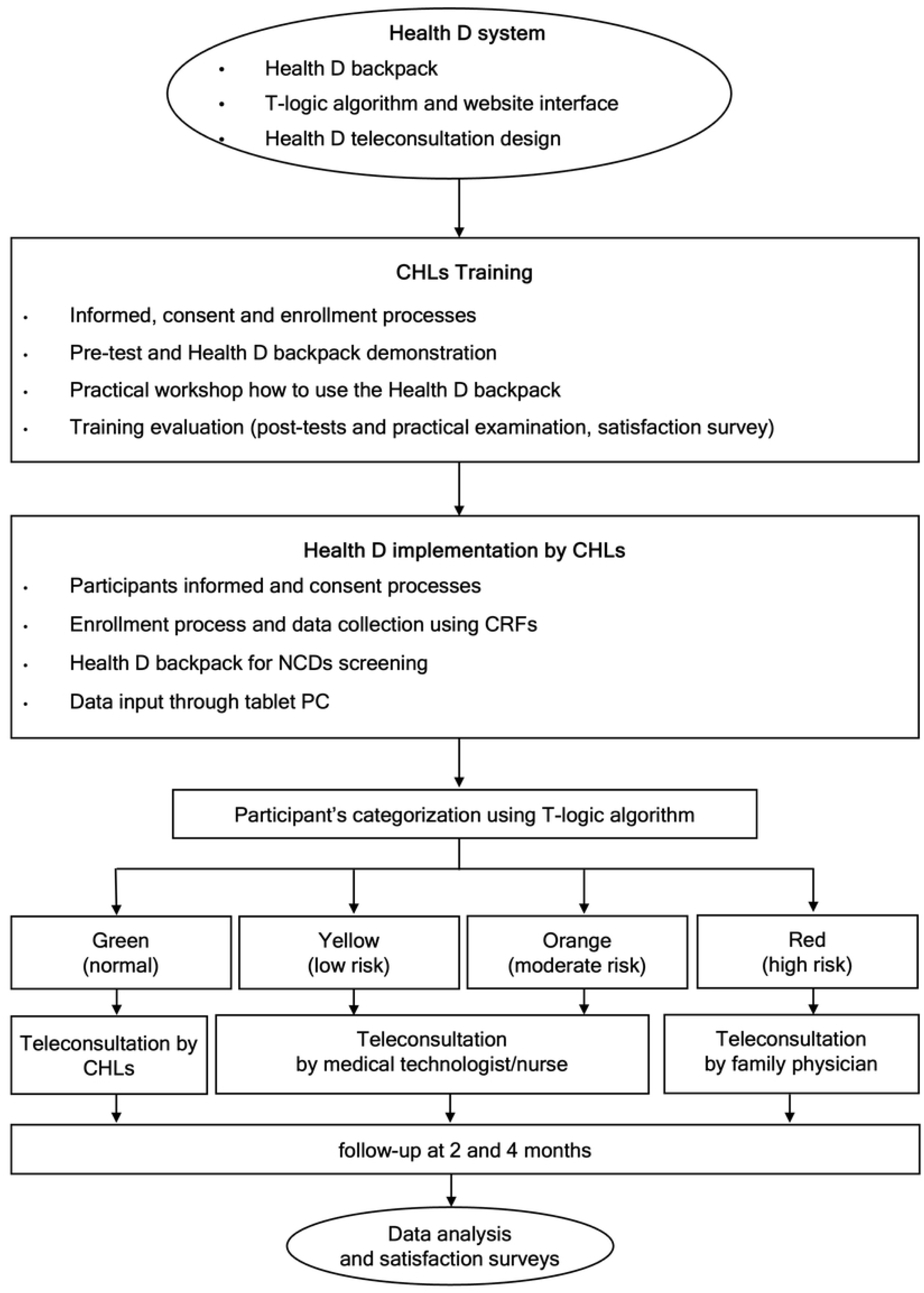
Framework of HealthD system.

### Preparation of the *HealthD* backpack

The *HealthD* backpack is equipped with a range of Thai Food and Drug Administration (FDA)-approved medical devices, ensuring accuracy and reliability in NCD screening and monitoring. It includes a digital thermometer (Omron MC-246), fingertip pulse oximeter (Yuwell YX102), and electronic blood pressure monitor (Yuwell YE670D) for routine vital sign assessments. Respiratory rate measurements were conducted manually. For physical examinations, the backpack contains a weighing scale (Xiaomi Smart Scale 2) and measurement tapes to assess height, weight, and waist-to-hip ratio (WHR). Additionally, the 6-minute walk test (6MWT) was used to evaluate participants’ aerobic capacity and functional endurance. Point-of-care testing (POCT) devices included a blood glucose monitor (ACCU-CHEK^®^ Instant), a lipid analyzer (Accutrend^®^ Plus), and a semi-quantitative urine analysis kit (Combur3 Test^®^), all validated for clinical reliability and diagnostic accuracy [23]. Additional essential equipment comprised sterile lancing devices, sanitary items, biohazard disposal tools, and a tablet PC for real-time data entry and teleconsultation, Fig 2.

**Fig 2.**
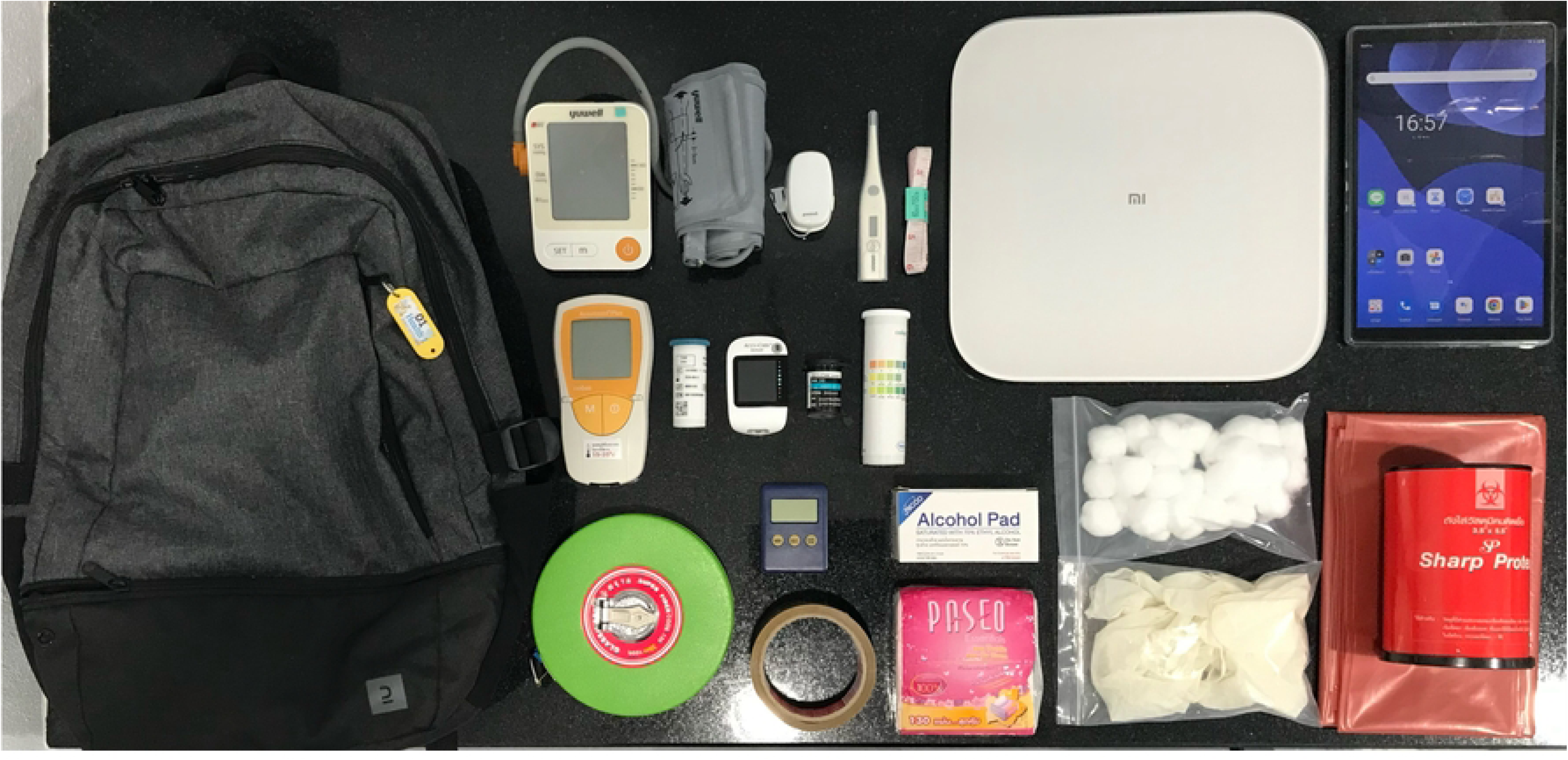
*HealthD* backpack and medical equipment.

### T-logic algorithm and website interface development

The T-logic algorithm developed using Visual Studio Code (v1.87) in JavaScript, classifies NCD risk levels based on international standards and Thai-specific clinical guidelines [24–26]. To ensure accessibility and ease of use, a user-friendly web interface was designed using HTML5 and CSS, enabling seamless data entry via tablet PCs, personal computers, and smartphones. For data storage and management, the system utilizes phpMyAdmin (v4.9.5) as the database management tool, while XAMPP (v8.2.12) was implemented to simulate data communication and optimize system performance before deployment. These features ensure efficient data handling, secure storage, and real-time accessibility, enhancing the overall functionality and reliability of the *HealthD* system, Fig 3.

**Fig 3.**
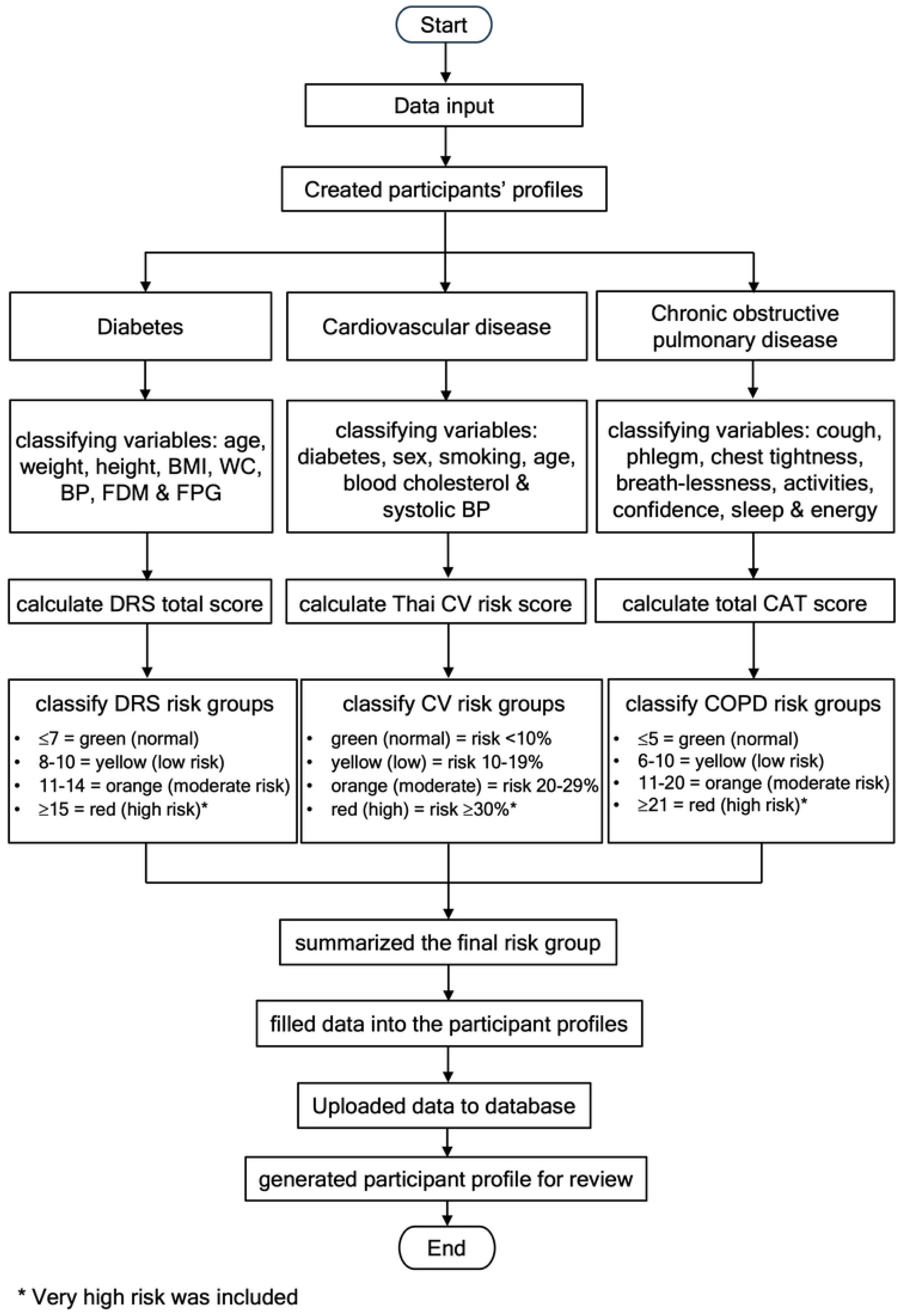
Framework of T-logic algorithm. BP: Blood pressure, BMI: Body Mass Index, CAT score: COPD Assessment Test score, DRS: Diabetes Risk Score, FPG: Fasting plasma glucose, FDM: Family history of Diabetes mellitus, Thai CV risk score: Thai Cardiovascular Risk Score, WC: Waist circumference

*HealthD* **teleconsultation design**

The *HealthD* teleconsultation framework integrates mobile messaging (Line^®^), tablet PCs, and smartphones to facilitate remote consultations between participants and healthcare professionals. The T-logic algorithm categorizes participants into four risk levels, directing “green” (normal-risk) cases to Community Health Leaders (CHLs) for general health guidance, while “yellow” (low-risk) and “orange” (moderate-risk) cases are referred to medical technologists or nurses for further assessment. Participants classified as “red” (high-risk) are escalated to family physicians for in-depth evaluation and medical intervention. The consultation process is guided by the Transtheoretical Model (Stages of Change), which supports behavioral modification strategies aimed at improving health outcomes and encouraging long-term adherence to preventive care [27].

### *HealthD* implementation

#### Study locations and ethical considerations

This study was conducted in Chiang Mai province, a mountainous region where access to healthcare remains a significant challenge. Specifically, the study focused on Doi Lo District, located approximately 45 km from Chiang Mai city, characterized by plateau and upland terrain at an average elevation of 300 meters [28]. The prospective pilot study was conducted between 24 June and 30 November 2023 across five rural villages: Don Chuen, Lao Pao, Lang Mon, Wang Kham Pom, and Huay Pao Yong. Ethical approval was obtained from the Research Ethics Committee (REC) of the Faculty of Associated Medical Sciences, Chiang Mai University (Approval Number:

AMSEC-66EX-011). Written informed consent was obtained from all participants before enrollment.

#### CHLs training

A one-day intensive training session was conducted to equip Community Health Leaders (CHLs) with the skills necessary to independently operate the *HealthD* system. CHLs with at least one month of prior experience in point-of-care testing (POCT) and proficiency in using tablet PCs were selected for participation. The training included a pre-training knowledge assessment, a comprehensive system overview, and hands-on demonstrations of the *HealthD* tools and software. CHLs engaged in guided practice sessions, followed by competency assessments to evaluate their technical proficiency. A post-training evaluation was conducted, requiring CHLs to achieve a minimum score of 80% to pass. At the conclusion of the training, participants completed a satisfaction survey to assess the acceptability of the program.

#### *HealthD* implementation by CHLs

Following training, each CHL was responsible for recruiting 10 participants, aged 30–70 years, with no prior diagnosis of NCDs, disabilities, or mental disorders. Written informed consent was obtained, and participants were assigned unique study codes to maintain confidentiality. CHLs collected comprehensive socio-demographic data, including medical history, vital signs, and physical examination results. NCD risk assessments were performed using the Diabetes Risk Score (DRS) for diabetes screening [24], the Thai Cardiovascular (Thai CV) Risk Score for CVD assessment [25], and the COPD Assessment Test (CAT) alongside the 6-Minute Walk Test (6MWT) for COPD risk stratification [26]. Data were recorded both on paper and in online case report forms (CRFs). Teleconsultations were conducted at two-month and four-month follow-ups, and at the conclusion of the study, participant satisfaction surveys were administered to evaluate the program’s acceptability.

### Statistical analysis

Baseline participant characteristics, including sex, age, occupation, income, education level, vital signs, height, weight, body mass index (BMI), waist-to-hip ratio (WHR), and NCD risk scores (diabetes, CVD, and COPD), were analyzed using categorical variables (expressed as numbers and percentages) and continuous variables (expressed as medians and interquartile ranges, IQR). CHLs’ pre- and post-training test scores were compared using the Wilcoxon signed-rank test, with statistical significance set at p *<*0.05 and a 95% confidence interval (CI). Satisfaction survey responses were evaluated using a 5-point Likert scale, with scores categorized as follows: 1.00–1.50 = Very dissatisfied, 1.51–2.50 = Dissatisfied, 2.51–3.50 = Neutral, 3.51–4.50 = Satisfied, 4.51–5.00 = Very satisfied. All statistical analyses were performed using Stata v16.0 (StataCorp, College Station, Texas).

## Results

### CHL competency assessment for ***HealthD*** backpack use

The competency of Community Health Leaders (CHLs) in operating the *HealthD* backpack was evaluated through pre- and post-training assessments. Among the 12 CHLs, 11 initially scored above 80%, with a mean pre-test score of 8.4 (SD = 0.9). Following the training, all CHLs achieved scores above 80%, with a significantly improved mean score of 9.3 (SD = 0.6). The observed increase in scores was statistically significant (*p* = 0.010), confirming that the training effectively enhanced CHLs’ knowledge and proficiency, ensuring their preparedness to implement the *HealthD* system in the field.

### Implementation of the ***HealthD*** system by CHLs

#### Socio-demographic characteristics of participants

A total of 120 participants were enrolled in this study, with a median age of 51 years (IQR: 43–59). The sample comprised 42 males (35%) and 78 females (65%). The majority of participants were freelance workers (60%), and 57% reported a monthly income ranging from 5,001 to 10,000 THB, while 31% earned below 5,000 THB. Regarding education levels, 54% had completed primary education, while 35% had attained secondary education, Table 1. Over the course of the study, one participant was lost to follow-up by the third visit.

**Table 1.** Socio-demographic characteristics of participants.

| Characteristics | All participants n=120 (%) |
| --- | --- |
| **Sex** |  |
| Male; n (%) | 42 (35) |
| Female; n (%) | 78 (65) |
| **Age median (IQR)** | 51 (43-59) |
| 30-40 years old; n (%) | 26 (22) |
| 41-50; n (%) | 34 (28) |
| 51-60; n (%) | 34 (28) |
| 61-70; n (%) | 26 (22) |
| **Occupation** |  |
| Agriculturist; n (%) | 14 (12) |
| Merchant; n (%) | 24 (20) |
| Company employee; n (%) | 3 (2) |
| Government officials & State enterprise employee; n (%) | 1 (1) |
| Freelance; n (%) | 72 (60) |
| Others; n (%) | 6 (5) |
| **Income (THB)** |  |
| <5,000; n (%) | 37 (31) |
| 5,001 - 10,000; n (%) | 68 (57) |
| 10,001 - 20,000; n (%) | 10 (8) |
| 20,001 - 30,000; n(%) | 3 (2) |
| >30,000; n (%) | 2 (2) |
| **Education levels** |  |
| <Primary school; n (%) | 4 (3) |
| Primary school; n (%) | 65 (54) |
| Secondary school; n (%) | 42 (35) |
| Bachelor's degree or equal; n (%) | 9 (8) |

#### Vital Signs

Vital signs data were recorded over three visits, revealing fluctuations in NCD risk classification. The normal-risk category accounted for 68%, 71%, and 66% of participants across the three visits, while the low-risk group comprised 24%, 23%, and 18%, respectively. The moderate-risk group constituted 7%, 5%, and 13%, whereas high-risk cases showed a slight increase from 1% at the first two visits to 3% at the final visit. Additionally, the prevalence of hypertension risk was 32%, 29%, and 34% across the three visits, Table 2.

**Table 2.** Results of *HealthD* implementation.

| Parameters | 1st visit (n=120) | 2nd visit (n=120) | 3rd visit (n=119) |
| --- | --- | --- | --- |
|  | n (%) or median (IQR) | n (%) or median (IQR) | n (%) or median (IQR) |
| Systolic blood pressure | 127 (116-138) | 128 (118-138) | 127 (118-140) |
| Diastolic blood pressure | 80 (76-88) | 81 (74-88) | 84 (75-90) |
| **Risk of hypertension** |  |  |  |
| Normal (110-139/80-89 mmHg) | 82 (68) | 85 (71) | 78 (66) |
| Low (140-159/90-99 mmHg) | 29 (24) | 28 (23) | 22 (18) |
| Moderate (160-179/100-109 mmHg) | 8 (7) | 6 (5) | 16 (13) |
| High ($\geq 180/\geq 110$ mmHg) | 1 (1) | 1 (1) | 3 (3) |
| **Body mass index (BMI), $kg/m^2$** | 24.2 (21.9-27.5) | 24.1 (22.0-27.5) | 24.3 (21.9-27.4) |
| **BMI groups** |  |  |  |
| Normal<br>(18.5-22.9 $kg/m^2$) | 41 (34) | 47 (39) | 48 (40) |
| Low<br>(23-24.9 $kg/m^2$) | 29 (24) | 22 (18) | 18 (15) |
| Moderate<br>(25-29.9 $kg/m^2$) | 32 (27) | 32 (27) | 34 (29) |
| High<br>($\geq 30.0$ $kg/m^2$) | 18 (15) | 19 (16) | 19 (16) |
| Waist-hip ratio (WHR) | 0.88 (0.84-0.92) | 0.88 (0.84-0.92) | 0.89 (0.84-0.91) |
| Male | 0.91 (0.86-0.94) | 0.91 (0.88-0.94) | 0.89 (0.87-0.95) |
| Female | 0.86 (0.80-0.92) | 0.86 (0.81-0.91) | 0.88 (0.83-0.91) |
| **Risk of obesity** |  |  |  |
| Normal | 54 (45) | 50 (42) | 52 (44) |
| Low | 33 (28) | 43 (36) | 32 (27) |
| Moderate | 23 (19) | 20 (17) | 24 (20) |
| High | 10 (8) | 7 (6) | 11 (9) |
| **Diabetes risk (DMR score)** | 10 (7-14) | 10 (6-13) | 11 (8-15) |
| Normal ($\leq 7$) | 32 (27) | 44 (37) | 29 (24) |
| Low (8-10) | 34 (28) | 26 (21) | 19 (16) |
| Moderate (11-14) | 29 (24) | 31 (26) | 40 (34) |
| High ($\geq 15$) | 25 (21) | 19 (16) | 31 (26) |
| **Cardiovascular risk (Thai CV risk)y** |  |  |  |
| Normal | 117 (97) | 115 (96) | 112 (94) |
| Low | 2 (2) | 5 (4) | 7 (6) |
| Moderate | 1 (1) | 0 (0) | 0 (0) |
| High | 0 (0) | 0 (0) | 0 (0) |
| Total cholesterol | 192 (173-218) | 193 (170-223) | 204 (177-229) |
| **COPD risk (CAT score)** |  |  |  |
| Normal ($\leq 5$) | 109 (90) | 113 (94) | 110 (92) |
| Low (6-10) | 9 (8) | 3 (3) | 9 (8) |
| Moderate (10-20) | 2 (2) | 4 (3) | 0 (0) |
| High (21-40) | 0 (0) | 0 (0) | 0 (0) |
| **6-minute walk test** |  |  |  |
| distance (m) | 455 (405-492) | 448 (420-480)<sup>a</sup> | 476 (440-507)<sup>b</sup> |
| **NCD risk groups** |  |  |  |
| Normal | 26 (22) | 41 (34) | 24 (20) |
| Low | 37 (31) | 26 (22) | 24 (20) |
| Moderate | 32 (26) | 34 (28) | 40 (34) |
| High | 25 (21) | 19 (16) | 31 (26) |

<sup>a</sup>Remained participants (n=117) for 6MWT, <sup>b</sup>Remained participants (n=116) for 6MWT

#### Physical Examination

The median body mass index (BMI) across visits was 24.2, 24.1, and 24.3 kg/m², classifying most participants as overweight based on Asian standards. The prevalence of overweight individuals was 66%, 61%, and 60% over the three visits. The waist-to-hip ratio (WHR), an indicator of abdominal obesity, remained high, with median values of 0.91 for males and 0.86 for females at the first visit, showing slight fluctuations in subsequent assessments. The prevalence of abdominal obesity remained consistently high at 55%, 58%, and 56% throughout the study period, Table 2.

### Screening for NCD Risks

#### Diabetes Risk

The median Diabetes Risk Score (DRS) was 10, 10, and 11 across the three visits. At baseline, 27% of participants were classified as normal risk, while 28% were low risk, 24% moderate risk, and 21% high risk. By the final visit, the proportions of moderate- and high-risk participants had increased to 34% and 26%, respectively, indicating a worsening trend in diabetes risk among some individuals, Table 2.

#### Cardiovascular Disease (CVD) Risk

At baseline, 97% of participants were classified as normal risk, with 2% and 1% categorized as low and moderate risk, respectively. By the third visit, the normal-risk proportion decreased slightly to 94%, while the low-risk category increased to 6%, reflecting minor fluctuations in CVD risk distribution, Table 2. ***COPD Risk:*** COPD risk, assessed using the COPD Assessment Test (CAT), indicated that the majority of participants maintained normal respiratory health, with 90%, 94%, and 92% classified as normal risk over the three visits. No participants were categorized as high-risk throughout the study. The 6-minute walk test (6MWT) results showed median distances of 455 m, 448 m, and 476 m across visits, remaining within the normal range for COPD assessment, Table 2. Correlation between 6MWT and CAT score for COPD risk assessment was a very weak negative correlation (*r* = -0.1111, *p* = 0.037) observed using linear regression, S1 Fig.

#### Overall NCD Risk Assessment

The overall NCD risk classification, integrating diabetes, CVD, and COPD assessments, revealed notable changes over time. At baseline, participants were classified as 22% normal risk, 31% low risk, 26% moderate risk, and 21% high risk. By the final visit, the proportions of normal- and low-risk participants decreased to 20%, while those categorized as moderate- and high-risk increased to 34% and 26%, respectively. This trend suggests that 66–80% of participants required continued teleconsultation support due to elevated NCD risk, Table 2. Risk progression was tracked across visits, S2 Fig, S3 Fig, and S4 Fig. Among the 26 participants initially classified as normal risk, 20 remained normal at the second visit, but this number declined to 14 by the final visit, with some shifting to higher-risk groups. In the low-risk category (37 participants initially), most either maintained or improved their risk status, although a few transitioned to moderate- or high-risk classifications. Among the 32 participants in the moderate-risk category at baseline, 14 remained stable, while others improved or regressed by the third visit. In the high-risk group (25 participants at baseline), approximately half remained in this category, while others improved temporarily before returning to high-risk status by the final visit, Fig 4.

**Fig 4.**
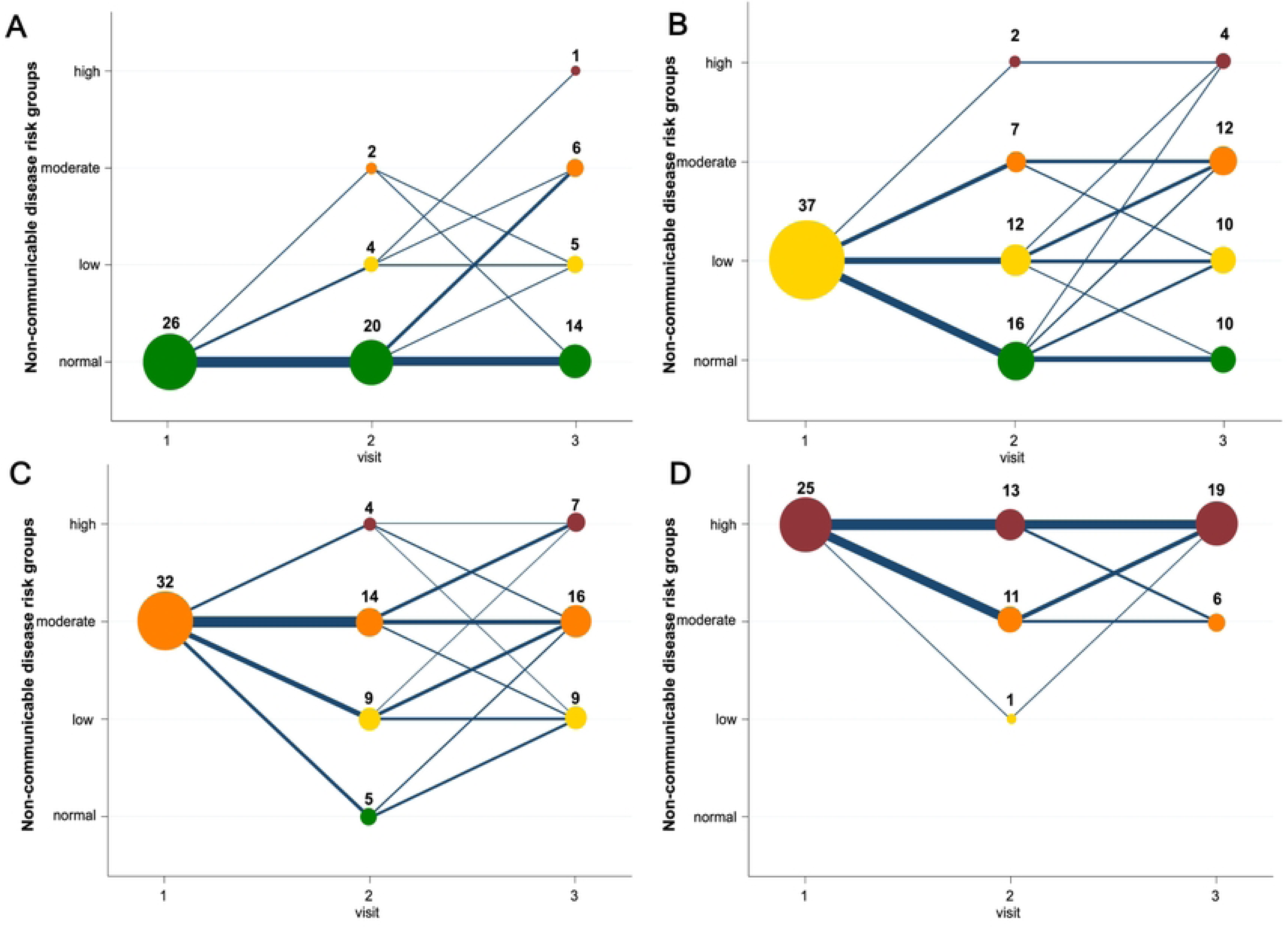
The number of participants being monitored for NCDs across three visits. A: normal group. B: low-risk group. C: moderate-risk group. D: high-risk group.

### Satisfaction Surveys of the ***HealthD*** System

#### Satisfactions of CHL Training and the ***HealthD*** Implementation

Satisfaction surveys were conducted to evaluate Community Health Leaders (CHLs)’ training experience and their perception of the *HealthD* system’s implementation. Overall, all CHLs reported satisfaction with the training, with an average session satisfaction score of at least 4.7 out of 5. Among the training components, 83% of CHLs expressed high satisfaction with modules covering vital signs assessment, physical examinations, POCT device demonstrations, presentation media, software interface usability, and overall program implementation. The COPD assessment module and training duration were particularly well-received, with 67% and 75% of CHLs rating them very highly, respectively Fig 5A. By the study’s conclusion, 92% of CHLs were satisfied with the overall HealthD implementation process, with an average satisfaction score of 4.6 out of 5. Only one CHL provided neutral feedback regarding teleconsultations, while the remaining ten CHLs expressed very high satisfaction with the system usability, and feasibility, reinforcing its potential for broader community-level implementation, Fig 5B.

**Fig 5.**
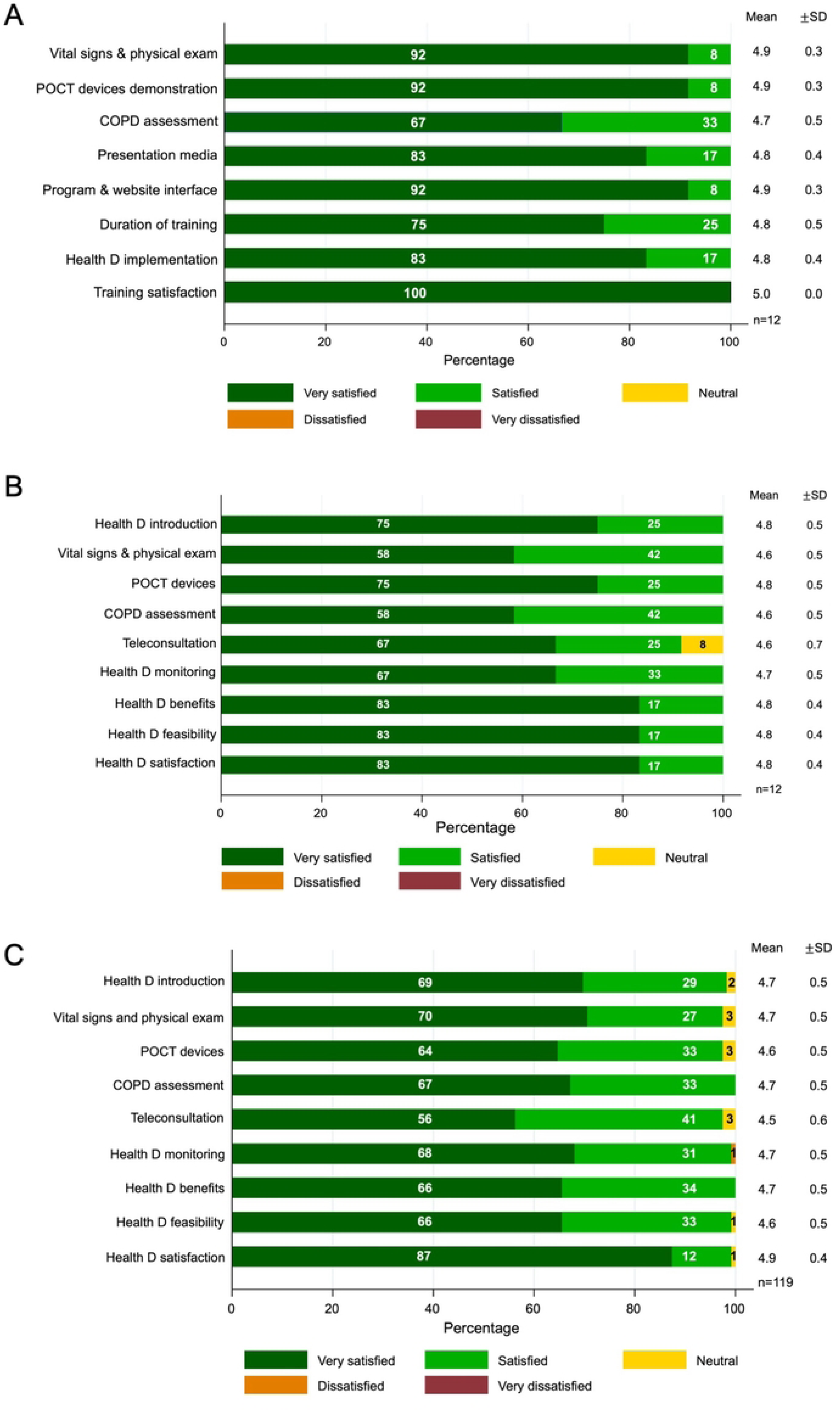
The proportion of *HealthD* satisfaction surveys obtained from stakeholders. A: The proportion of CHLs training satisfaction surveys (n=12). B: The proportion of the *HealthD* system satisfaction surveys obtained from the CHLs (n=12). C: The proportion of the *HealthD* system satisfaction surveys obtained from the participants (n=119)

#### Participant Satisfaction

The participant satisfaction survey indicated that 97% of participants (n = 116) were satisfied with all aspects of the *HealthD* system, with an average satisfaction rating of at least 4.5 out of 5. More than half (56%) reported being very satisfied, underscoring the system’s acceptability and ease of use. However, one participant expressed dissatisfaction with the *HealthD* monitoring process. Despite this, the perceived value of the *HealthD* system remained overwhelmingly positive, with 99% of participants believing in its feasibility and sustainable integration within their community, Fig 5C.

## Discussion

This study evaluated the feasibility of the *HealthD* system, a telehealth platform designed for community-based screening and monitoring of non-communicable diseases (NCDs) in Doi Lo District, Chiang Mai, Thailand. Operated by trained Community Health Leaders (CHLs), the system facilitated vital sign monitoring, physical examinations, and biomarker analysis, categorizing participants into NCD risk groups and providing teleconsultations based on their risk level. The high satisfaction rates among both CHLs and participants suggest that the *HealthD* system is well-accepted and feasible for community implementation, contributing to enhanced health awareness and early disease detection in underserved areas. The *HealthD* system integrates validated biomedical tools and the T-logic algorithm, which is adapted to the Thai population while adhering to international standards. Similar models, such as Bangladesh’s B-logic system in the Portable Health Clinic (PHC) box [29], have demonstrated effectiveness in remote NCD monitoring across various settings, including unreached communities, maternal and childcare programs, and NCD screening in Indonesia [13–16]. During the COVID-19 pandemic, the PHC model was successfully adapted for infectious disease screening, showcasing its flexibility in resource-limited settings [17, 30]. The successful integration of *HealthD* in rural Thailand reinforces its potential as a scalable telehealth solution for addressing healthcare accessibility challenges in underserved populations. The *HealthD* system’s web-based interface supports multi-device access, emphasizing ease of use and scalability. This approach aligns with other digital health interventions, such as Hong Kong’s “Perfect Diet and Exercise” platform for older adults [31], India’s “One Health” mobile app for hypertension management [18], and asthma self-management applications for adolescents [32]. Studies indicate that web-based and mobile health applications significantly enhance NCD monitoring, though technological literacy remains a critical factor, particularly among older adults [31]. In this study, CHLs played a central role in bridging the digital divide, providing direct assistance to participants and facilitating effective system adoption. The effectiveness of CHL-led health interventions is well-documented. CHL training significantly improved competency levels, mirroring results from peer-training programs in Thailand [33] and India’s telementoring model for primary healthcare workers [34]. Moreover, WHO reports confirm that well-trained community health workers can effectively perform NCD screenings and data collection, emphasizing the importance of continuous training and refresher courses to maintain long-term effectiveness [35]. Hypertension and diabetes were primary concerns in this study. Although average blood pressure remained within normal ranges, one-third of participants were classified as hypertensive, exceeding the national prevalence of 25.4% reported in the 6th Thailand National Health Examination Survey (NHES-VI) [36]. Additionally, two-thirds of participants were overweight, a figure comparable to national obesity rates, with low income and education levels contributing to poor dietary habits and sedentary lifestyles [37–39]. Diabetes screening indicated that 75% of participants were at risk, with risk distributions remaining relatively stable, though some participants improved their behaviors over time. Cultural dietary factors, such as the prevalence of high-carbohydrate foods like sticky rice and sweet fruits, likely contributed to elevated diabetes risk in the study population [40]. CVD risk assessments remained stable, though the short study duration may have limited the ability to detect significant trends. Meanwhile, COPD risk assessments showed that most participants were classified as normal risk, but future risks may increase due to regional air pollution exposure. PM2.5 pollution in Chiang Mai has been linked to higher COPD-related mortality and hospitalizations, highlighting the need for long-term respiratory health monitoring [41, 42]. Interestingly, the study found a weak negative correlation between CAT scores and 6MWT distance, contrasting prior studies that observed stronger correlations in COPD patients [43]. Factors such as age, sex, BMI, and variations in testing environments may have influenced results [44–46], while familiarity with the 6MWT appeared to improve participant performance over time [47]. Although the 6MWT is a cost-effective and practical assessment tool, its reliability for COPD risk assessment could be enhanced by integrating spirometry in future screenings. The combined NCD risk assessment revealed that while CVD and COPD risks remained relatively stable, diabetes risk showed fluctuations. A temporary improvement was noted at the second visit, but the final assessment indicated a rise in risk levels, potentially due to the Hawthorne effect, where participants initially alter behaviors under observation but gradually revert to prior habits [48]. Compared to similar studies, HealthD participants exhibited higher NCD risk levels, underscoring the need for continuous primary care interventions and community-based health promotion [13, 15]. Teleconsultations within the *HealthD* system were well-received, particularly during follow-up visits, where healthcare professionals provided personalized guidance. This model not only reduces hospital visits but also enhances patient engagement and lowers healthcare costs. However, participant willingness to engage with teleconsultations is crucial for success. Previous studies indicate that younger, tech-savvy individuals are more receptive to telehealth, while older adults often require family support [49–51]. To accommodate diverse technological capabilities, the *HealthD* system also offered telephone support, ensuring accessibility for participants uncomfortable with video calls. Overall, both CHLs and participants reported high satisfaction with the *HealthD* system. CHLs valued the training program, and 97% of participants expressed satisfaction with the system’s practicality and health benefits. However, missed teleconsultation appointments were noted as a challenge, highlighting the need for more flexible scheduling options. A comparable study in Bahrain reported a 92% telehealth satisfaction rate, aligning with *HealthD*’s positive reception and potential scalability [52]. Several limitations should be acknowledged. The small sample size and gender imbalance (higher proportion of female participants) may limit generalizability. Additionally, the two-month interval between follow-up visits resulted in some CHLs requiring refresher training for POCT devices. Internet connectivity issues occasionally interfered with real-time teleconsultations, impacting user experience. Furthermore, the exclusion of cancer markers highlights the need for a more comprehensive NCD screening approach. Future research should explore the long-term impact of the *HealthD* system on NCD management, including behavioral changes, adherence to lifestyle modifications, and teleconsultation effectiveness. Expanding the study to larger populations and diverse geographical settings would provide greater insight into its scalability.

## Conclusion

In conclusion, the *HealthD* system demonstrated strong feasibility for telehealth-based NCD screening and monitoring in rural Thailand. By enhancing healthcare access, improving early disease detection, and fostering community-driven health engagement, the platform holds significant potential for scaling up in resource-limited settings. The high satisfaction rates among CHLs and participants underscore its practicality and acceptability. With ongoing telehealth investment and refinement, the *HealthD* system could play a pivotal role in strengthening NCD prevention and management, particularly in underserved communities.

## Supporting information

**S1 Fig. Correlation between CAT score and 6-minute walk distance (6MWD).** *r* = -0.1111, *p* = 0.037).

**S2 Fig. The number of participants being monitored for diabetes across three visits categorized by diabetes risk score (A-D).** A: normal group. B: low-risk group. C: moderate-risk group. D: high-risk group.

**S3 Fig. The number of participants being monitored for CVD across three visits categorized by Thai CV risk score.** A: normal group. B: low-risk group. C: moderate-risk group.

**S4 Fig. The number of participants being monitored for COPD across three visits categorized by CAT score.** A: normal group. B: low-risk group. C: moderate-risk group.

## Data Availability

All relevant data are within the manuscript and its Supporting Information files.

## Acknowledgments

We extend our gratitude to the Community Health Leaders (CHLs) and all participants for their valuable contributions to this study. We also sincerely appreciate the Chief Executive of the Doi Lo Subdistrict Administrative Organization for their facilitation and support.

## References

1. World Health Organization. Noncommunicable diseases Geneva;2022 [updated 2022 Sep 16; cited 2022 Nov 27]. Available from: https://www.who.int/news-room/fact-sheets/detail/noncommunicable-diseases.

2. World Health Organization. Noncommunicable Diseases Progress Monitor 2022 Geneva;2022 [cited 2022 Nov 27]. Available from: https://iris.who.int/bitstream/handle/10665/353048/9789240047761-eng.pdf?sequence=1&isAllowed=y.

3. World Health Organization. NCD Data Portal Geneva;2019 [cited 2022 Dec 31]. Available from: https://ncdportal.org/CountryProfile/GHE110/Thailand.

4. Ministry of Public Health, Division of Non Communicable Disease. Number and mortality rate from 5 non-communicable diseases in 2018 - 2022 Muang Nonthaburi;2022 [updated 2023 Nov 1; cited Aug 2023 21]. Available from: https://www.ddc.moph.go.th/dncd/news.php?news=39911.

5. Boonma A, Sethanan K, Talangkun S, Laonapakul T. Patient waiting time and satisfaction in GP clinic at a tertiary hospital in Thailand. MATEC Web of Conferences. 2018;192:01034. doi: 10.1051/matecconf/201819201034.

6. World Health Organization. Prevention and control of noncommunicable diseases in Thailand : The case for investment Geneva;2021 [cited 2022 Nov 23]. Available from: https://thailand.un.org/en/159788-prevention-and-control-noncommunicable-diseases-thailand-case-investment.

7. Yingtaweesak T, Yoshida Y, Hemhongsa P, Hamajima N, Chaiyakae S. Accessibility of health care service in Thasongyang, Tak Province, Thailand. Nagoya J Med Sci. 2013;75(3-4):243–50. doi: PMC4345673.

8. Khemapech I. Telemedicine – Meaning, Challenges and Opportunities. Siriraj Medical Journal. 2019;71:246–52. doi: 10.33192/Smj.2019.38.

9. Sudhipongpracha T, Poocharoen O-O. Community Health Workers as Street-level Quasi-Bureaucrats in the COVID-19 Pandemic: The Cases of Kenya and Thailand. Journal of Comparative Policy Analysis: Research and Practice. 2021;23(2):234–49. doi: 10.1080/13876988.2021.1879599.

10. Aubrecht P, Essink J, Kovac M, Vandenberghe A-S. Centralized and decentralized responses to COVID-19 in federal systems: US and EU comparisons. SSRN 3584182. 2020. doi: 10.2139/ssrn.3584182.

11. Scott K, Beckham SW, Gross M, Pariyo G, Rao KD, Cometto G, et al. What do we know about community-based health worker programs? A systematic review of existing reviews on community health workers. Hum Resour Health. 2018;16(1):39. doi: 10.1186/s12960-018-0304-x.

12. World Health Organization and International Telecommunication Union 2022. WHO-ITU global standard for accessibility of telehealth services Geneva;2022 [updated 2022 Jan 1; cited 2023 Jan 10]. Available from: https://iris.who.int/bitstream/handle/10665/356160/9789240050464-eng.pdf?sequence=1.

13. Islam R, Nohara Y, Rahman MJ, Sultana N, Ahmed A, Nakashima N. Portable Health Clinic: An Advanced Tele-Healthcare System for Unreached Communities. Stud Health Technol Inform. 2019;264:616–9. doi: 10.3233/shti190296.

14. Ahmed A, Inoue S, Kai E, Nakashima N, Nohara Y. Portable Health Clinic: A Pervasive Way to Serve the Unreached Community for Preventive Healthcare. 2013;8028:265–74. doi: 10.1007/978-3-642-39351-8_29.

15. Wulandari H, Lazuardi L, Majid N, Yokota F, Sanjaya GY, Dewi TS, et al. Potential Improvement in a Portable Health Clinic for Community Health Service to Control Non-Communicable Diseases in Indonesia. Applied Sciences. 2023;13(3):1623. doi: 10.3390/app13031623.

16. Islam R, Kikuchi K, Sato Y, Izukura R, Jahan N, Sultana N, et al. Maternal and Child Healthcare Service by Portable Health Clinic System Using a Triage Protocol. Stud Health Technol Inform. 2021;284:130–4. doi: 10.3233/shti210684.

17. Islam R, Yokota F, Nishikitani M, Kikuchi K, Sato Y, Izukura R, et al. Portable health clinic COVID-19 system for remote patient follow-up ensuring clinical safety. Comput Methods Programs Biomed Update. 2022;2:100061. doi: 10.1016/j.cmpbup.2022.100061.

18. S AK, Bali S, Pakhare AP, Khadanga S. Feasibility of Self-Management of Hypertension and Diabetes Using Patient-Generated Health Data Through M-health in Central India. Cureus. 2024;16(2):e55060. doi: 10.7759/cureus.55060.

19. Sirintrapun SJ, Lopez AM. Telemedicine in Cancer Care. Am Soc Clin Oncol Educ Book. 2018;38:540–5. doi: 10.1200/edbk_200141.

20. Amatavete S, Lujintanon S, Teeratakulpisarn N, Thitipatarakorn S, Seekaew P, Hanaree C, et al. Evaluation of the integration of telehealth into the same-day antiretroviral therapy initiation service in Bangkok, Thailand in response to COVID-19: a mixed-method analysis of real-world data. J Int AIDS Soc. 2021;24 Suppl 6:e25816. doi: 10.1002/jia2.25816.

21. Mamom J, Daovisan H. Telenursing: How do caregivers treat and prevent pressure injury in bedridden patients during the COVID-19 pandemic in Thailand? Using an embedded approach. J Telemed Telecare. 2022 doi: 10.1177/1357633x221078485.

22. Stonsaovapak C, Sangveraphunsiri V, Jitpugdee W, Piravej K. Telerehabilitation in Older Thai Community-Dwelling Adults. Life (Basel). 2022;12(12):2029–40. doi: 10.3390/life12122029.

23. Maruf RI, Tou S, Izukura R, Sato Y, Nishikitani M, Kikuchi K, et al. An evaluation of the commonly used portable medical sensors performance in comparison to clinical test results for telehealth systems. Computer Methods and Programs in Biomedicine Update. 2024;5:100147. doi: 10.1016/j.cmpbup.2024.100147.

24. Diabetes Association of Thailand under The Patronage of Her Royal Highness Princess Maha Chakri Sirindhorn, The Endocrine Society of Thailand. Clinical Practice Guideline for Diabetes 2023. Bangkok: 2023 [cited 2023 1 NOV]. Available from: https://www.thaiendocrine.org/th/wp-content/uploads/2023/08/Thai-DM-CPG-2566.pdf.

25. Ministry of Public Health, Department of Disease Control, Division of Non Communicable Disease. Handbook of Cardiovascular Disease Assessment for Village Health Volunteers (VHV). Muang Nonthaburi; 2016.

26. Jones PW, Harding G, Berry P, Wiklund I, Chen WH, Kline Leidy N. Development and first validation of the COPD Assessment Test. Eur Respir J. 2009;34(3):648–654. doi: 10.1183/09031936.00102509.

27. Wayne W. LaMorte M, PhD, MPH. The Transtheoretical Model (Stages of Change): Boston University School of Public Health; 2022 [updated 2022 Nov 03; cited 2024 Apr 09]. Available from: https://sphweb.bumc.bu.edu/otlt/mph-modules/sb/behavioralchangetheories/behavioralchangetheories6.html.

28. Doi Lo Subdistrict Administrative Organization. History and information of Doi Lo Subdistrict Administrative Organization Chiang Mai;2006 [cited 2023 Jan 3]. Available from: http://www.doilor.org/about.php?id=1.

29. Ahmed A, Rebeiro-Hargrave A, Nohara Y, Kai E, Ripon Z, Nakashima N. Targeting Morbidity in Unreached Communities Using Portable Health Clinic System. IEICE Trans Commun. 2014;E97-B:540-45. doi: 10.1587/transcom.E97.B.540.

30. Sampa MB, Hoque MR, Islam R, Nishikitani M, Nakashima N, Yokota F, et al. Redesigning Portable Health Clinic Platform as a Remote Healthcare System to Tackle COVID-19 Pandemic Situation in Unreached Communities. Int J Environ Res Public Health. 2020;17(13): doi: 10.3390/ijerph17134709.

31. Yang M, Duan Y, Lippke S, Liang W, Su N. A blended face-to-face and eHealth lifestyle intervention on physical activity, diet, and health outcomes in Hong Kong community-dwelling older adults: a study protocol for a randomized controlled trial. Front Public Health. 2024;12:1360037. doi: 10.3389/fpubh.2024.1360037.

32. Carpenter DM, Geryk LL, Sage A, Arrindell C, Sleath BL. Exploring the theoretical pathways through which asthma app features can promote adolescent self-management. Transl Behav Med. 2016;6(4):509–18. doi: 10.1007/s13142-016-0402-z.

33. Pitchalard K, Moonpanane K, Wimolphan P, Singkhorn O, Wongsuraprakit S. Implementation and evaluation of the peer-training program for village health volunteers to improve chronic disease management among older adults in rural Thailand. Int J Nurs Sci. 2022;9(3):328–33. doi: 10.1016/j.ijnss.2022.06.011.

34. Zaman SB, Evans RG, Singh R, Singh A, Singh P, Singh R, et al. Feasibility of community health workers using a clinical decision support system to screen and monitor non-communicable diseases in resource-poor settings: study protocol. Mhealth. 2021;7:15. doi: 10.21037/mhealth-19-258.

35. World Health Organization, Regional Office for South-East Asia. Use of community health workers to manage and prevent noncommunicable diseases: policy options based on the findings of the COACH study. India, 2019 [cited 2024 Jul 09]. Available from: https://iris.who.int/handle/10665/325733.

36. Wichai Aekplakorn, Hataichanok Puckcharern, Warapone Satheannoppakao. The 6th Thailand National Health Examination Survey (NHES-VI) 2019-2020.: Aksorn graphic and design publishing limited partnershipk; 2021 [cited 2024 Aug 23].

37. Phulkerd S, Thongcharoenchupong N, Chamratrithirong A, Gray RS, Pattaravanich U, Ungchusak C, et al. Socio-demographic and geographic disparities of population-level food insecurity during the COVID-19 pandemic in Thailand. Front Public Health. 2022;10:1071814. doi: 10.3389/fpubh.2022.1071814.

38. Mambrini SP, Menichetti F, Ravella S, Pellizzari M, De Amicis R, Foppiani A, et al. Ultra-Processed Food Consumption and Incidence of Obesity and Cardiometabolic Risk Factors in Adults: A Systematic Review of Prospective Studies. Nutrients. 2023;15(11):doi: 10.3390/nu15112583.

39. Biswas T, Townsend N, Huda MM, Maravilla J, Begum T, Pervin S, et al. Prevalence of multiple non-communicable diseases risk factors among adolescents in 140 countries: A population-based study. EClinicalMedicine. 2022;52:101591. doi: 10.1016/j.eclinm.2022.101591.

40. Yeemard F, Srichan P, Apidechkul T, Luerueang N, Tamornpark R, Utsaha S. Prevalence and predictors of suboptimal glycemic control among patients with type 2 diabetes mellitus in northern Thailand: A hospital-based cross-sectional control study. PLoS One. 2022;17(1):e0262714. doi: 10.1371/journal.pone.0262714.

41. Sapbamrer P, Assavanopakun P, Panumasvivat J. Decadal Trends in Ambient Air Pollutants and Their Association with COPD and Lung Cancer in Upper Northern Thailand: 2013-2022. Toxics. 2024;12(5): doi: 10.3390/toxics12050321.

42. Pipatvech K, Uppachak S. Relationship between PM2.5 Dust Exposure and Exacerbation of Chronic Obstructive Pulmonary Disease Patients in Nan Hospital, Thailand. Journal of Health Science of Thailand. 2021;30(4):645–53.

43. Konjeti O, Gaikwad N, Manjush R, Saini M. Correlation of Six-Minute Walk Test and COPD Assessment Test (CAT) Scores with Spirometric Indices in Chronic Obstructive Pulmonary Disease Patients. International Journal of Medical Reviews and Case Reports. 2022;6(14):24–28. doi: 10.5455/IJMRCR.172-1646754928.

44. Kim AL, Kwon JC, Park I, Kim JN, Kim JM, Jeong BN, et al. Reference equations for the six-minute walk distance in healthy korean adults, aged 22-59 years. Tuberc Respir Dis (Seoul). 2014;76(6):269–75. doi: 10.4046/trd.2014.76.6.269.

45. Cazzoletti L, Zanolin ME, Dorelli G, Ferrari P, Dalle Carbonare LG, Crisafulli E, et al. Six-minute walk distance in healthy subjects: reference standards from a general population sample. Respir Res. 2022;23(1):83. doi: 10.1186/s12931-022-02003-y.

46. Zou H, Zhu X, Zhang J, Wang Y, Wu X, Liu F, et al. Reference equations for the six-minute walk distance in the healthy Chinese population aged 18-59 years. PLoS One. 2017;12(9):e0184669. doi: 10.1371/journal.pone.0184669.

47. Agarwala P, Salzman SH. Six-Minute Walk Test: Clinical Role, Technique, Coding, and Reimbursement. Chest. 2020;157(3):603–11. doi: 10.1016/j.chest.2019.10.014.

48. Bootwong P, Intarut N. The Effects of Text Messages for Promoting Physical Activities in Prediabetes: A Randomized Controlled Trial. Telemed J E Health. 2022;28(6):896–903. doi: 10.1089/tmj.2021.0303.

49. Orrange S, Patel A, Mack WJ, Cassetta J. Patient Satisfaction and Trust in Telemedicine During the COVID-19 Pandemic: Retrospective Observational Study. JMIR Hum Factors. 2021;8(2):e28589. doi: 10.2196/28589.

50. Sin DYE, Guo X, Yong DWW, Qiu TY, Moey PKS, Falk MR, et al. Assessment of willingness to Tele-monitoring interventions in patients with type 2 diabetes and/or hypertension in the public primary healthcare setting. BMC Med Inform Decis Mak. 2020;20(1):11. doi: 10.1186/s12911-020-1024-4.

51. Rockler Meurling C, Adell E, Wolff M, Calling S, Milos Nymberg V, Borgström Bolmsjö B. Telemedicine in Swedish primary health care - a web-based survey exploring patient satisfaction. BMC Health Serv Res. 2023;23(1):129. doi: 10.1186/s12913-023-09133-z.

52. Habbash F, Rabeeah A, Huwaidi Z, Abuobaidah H, Alqabbat J, Hayyan F, et al. Telemedicine in non-communicable chronic diseases care during the COVID-19 pandemic: exploring patients’ perspectives. Front Public Health. 2023;11:1270069. doi: 10.3389/fpubh.2023.1270069.

